# Impulsivity Facets and Substance Use Involvement: Insights from Genomic Structural Equation Modeling

**DOI:** 10.1101/2024.10.24.24315331

**Authors:** Laura Vilar-Ribó, Alexander S Hatoum, Andrew D Grotzinger, Travis T Mallard, 23andMe Research Team, Sarah Elson, Pierre Fontanillas, Abraham A Palmer, Daniel E Gustavson, Sandra Sanchez-Roige

## Abstract

**Background:** Impulsivity is a multidimensional trait associated with substance use disorders (SUDs), but the relationship between distinct impulsivity facets and stages of substance use involvement remains unclear.

**Methods:** We used genomic structural equation modeling and genome-wide association studies (N=79,729-903,147) based on individuals of European ancestry to examine the latent genetic architecture of impulsivity and substance use behaviors. We included GWAS data from nine impulsivity traits and seven substance use (SU) and SUD traits.

**Results:** We found that the SU and SUD factors were strongly genetically inter-correlated (*r_G_*=0.77) but their associations with impulsivity factors differed. Lack of premeditation, negative and positive urgency were equally positively genetically correlated with both the SU (*r_G_*=.0.30-0.50) and SUD (*r_G_=*0.37-0.47) factors; sensation seeking was more strongly genetically correlated with the SU factor (*r_G_*=0.27 vs. *r_G_*=0.10); delay discounting was more strongly genetically correlated with the SUD factor (*r_G_*=0.32 vs. *r_G_*=0.21); and lack of perseverance was only weakly genetically correlated with the SU factor (*r_G_*=0.09). After controlling for the genetic correlation between SU/SUD, we found that lack of premeditation and positive urgency were independently genetically associated with both the SU (β=0.39, β=0.28, respectively) and SUD factors (β=0.28, β=0.19, respectively); sensation seeking and lack of perseverance were independently genetically associated with the SU factor (β=0.46, β=0.19, respectively); and negative urgency and delay discounting were independently genetically associated with the SUD factor (β=0.34, β=0.39, respectively).

**Conclusions:** Our findings show that specific impulsivity facets confer risk for distinct stages of substance use involvement, with potential implications for SUDs prevention and treatment.

## Introduction

Impulsivity, broadly defined as the tendency to act on urges or desires without forethought or consideration of potential consequences, is a heritable trait associated with numerous psychiatric disorders, including substance use disorders (SUDs) (Amlung et al., 2017; Berg et al., 2015; Ersche et al., 2010; Kozak et al., 2019; Lees et al., 2021; MacKillop et al., 2011; Mitchell, 2019; Vassileva & Conrod, 2019). As the concept of impulsivity has evolved, researchers have recognized that it represents a family of cognitive and behavioral tendencies influenced by partially distinct neurobiological substrates (E. Levitt et al., 2020; Meda et al., 2009; Miller & Gizer, 2023; Vassileva & Conrod, 2019), as opposed to a single unitary construct (Dick et al., 2010; Evenden, 1999; Griffin et al., 2018). These different forms can manifest, for instance, through motor (acting without thinking), non-planning (lack of forethought), choice (preference for immediate rewards), risk-taking (engaging in potentially harmful activities) or attentional (difficulty focusing) impulsivity. To thoroughly assess this multi-dimensionality, several psychological instruments have been developed, including self-reported questionnaires such as the Impulsive Behavior Scale (UPPS-P) (Cyders et al., 2014; Whiteside et al., 2005), the Barratt Impulsiveness scale (BIS) (Barratt, 1994), and the Monetary Choice Questionnaire (MCQ) (Kirby et al., 1999).

Impulsivity is a well-established risk factor for SUDs, and is known to influence several stages of vulnerability, including initial substance use, regular use without consequences or dependence (“normative use”; Kearns et al., 2022; Shin et al., 2012; Wasserman et al., 2020, 2021), as well as the progression towards compulsive and problematic use (Lee et al., 2019; Petker et al., 2021; Poulton & Hester, 2020; Verdejo-Garcia & Albein-Urios, 2021), and treatment outcomes (Athamneh et al., 2019, 2022; Heinz et al., 2015; E. E. Levitt et al., 2023; Loree et al., 2015). In addition, these epidemiological studies have suggested that specific impulsivity facets have unique relationships with these different stages of substance use vulnerability. For instance, sensation seeking is often associated with substance use initiation, whereas other facets, such as negative urgency, lack of premeditation and delay discounting, are associated with substance use-related problems or poorer treatment outcomes (Hershberger et al., 2017; Hildebrandt et al., 2021; Kearns et al., 2022; Kräplin et al., 2020; MacKillop et al., 2011; Petker et al., 2021; Stamates & Lau-Barraco, 2017; Stautz & Cooper, 2013; Tran et al., 2018). A deeper knowledge into these differences could lead to more effective strategies for treating and preventing SUDs by targeting specific dimensions of impulsivity. However, these prior phenotypic studies often face methodological challenges, such as small sample sizes, variability in instrument measurement and difficulty controlling for potential environmental biases, which can limit the generalizability and interpretability of the findings (Friedman & Gustavson, 2022; Sanchez-Roige & Palmer, 2020).

Genetic studies can partially overcome some of the limitations of phenotypic studies by examining the relationship between impulsivity and substance use vulnerability using independent cohorts, which partially controls for potential environmental confounds (Miller & Gizer, 2024; Sanchez-Roige, Fontanillas, et al., 2019; Sanchez-Roige et al., 2023). For more than two decades, family and twin studies have shown that the relationships between impulsivity and substance use behaviors are largely due to an underlying shared genetic liability (Kendler et al., 2003; Slutske et al., 2002; Tarter et al., 2004). Recent multivariate techniques, such as Genomic Structural Equation Modeling (Genomic SEM), can combine data from multiple correlated phenotypes derived from genome-wide association studies (GWAS) and model the latent genetic factor structure of impulsivity and substance use behaviors (Grotzinger et al., 2019). Using recent GWAS of impulsivity (Sanchez-Roige, Fontanillas, et al., 2019; Sanchez-Roige et al., 2023), we have applied Genomic SEM to model the genetic architecture of impulsivity facets and corroborated that impulsivity is multidimensional (Gustavson et al., 2020, 2024; Mallard et al., 2023). While most impulsivity facets were positively genetically correlated with one another, others such as sensation seeking and delay discounting were divergent (Gustavson et al., 2020, 2024; Mallard et al., 2023). We also used Genomic SEM to model the latent structure of SUDs, and shown that the genetic architecture of normative use and SUDs are distinct (Hatoum et al., 2022, 2023; Mallard et al., 2022). Furthermore, a recent Genomic SEM study identified a stronger genetic overlap between sensation seeking and alcohol consumption compared to alcohol use disorder (Miller & Gizer, 2024). However, the extent to which other impulsivity facets are differentially genetically related to aspects of substance use involvement has not been systematically explored.

Here we build upon these Genomic SEM findings to model the genetic architecture of impulsivity facets and substance use behaviors. We leveraged access to well-powered GWAS summary statistics (N=79,729-903,147) to examine the latent genetic relationship between nine impulsivity facets, captured by six latent factors based on our prior work, including negative and positive urgency (inability to resist temptations while experiencing positive or negative affect), lack of premeditation (tendency to act without planning or self-control), lack of perseverance (inability to persist on difficult tasks), sensation seeking (tendency to enjoy risky situations) and delay discounting (preference for smaller immediate rewards over larger delayed rewards), and seven substance use-related measures, captured by two factors based on our prior work (Hatoum et al., 2022, 2023), including normative substance use (SU; based on lifetime and quantity/frequency use) and SUD. Based on prior findings, we hypothesized that specific impulsivity facets would be differentially associated with SU and SUD factors, providing a more comprehensive understanding into the etiology of substance use vulnerability.

## Methods

### GWAS data

#### Overview

We used published GWAS derived from genetically-predicted individuals of European ancestry based on genetic similarity (National Academies of Sciences & Medicine, 2023). Our study was limited to this group because similar impulsivity GWAS data are not available for other groups. **Table 1** provides an overview of the GWAS datasets used.

**Table 1.**
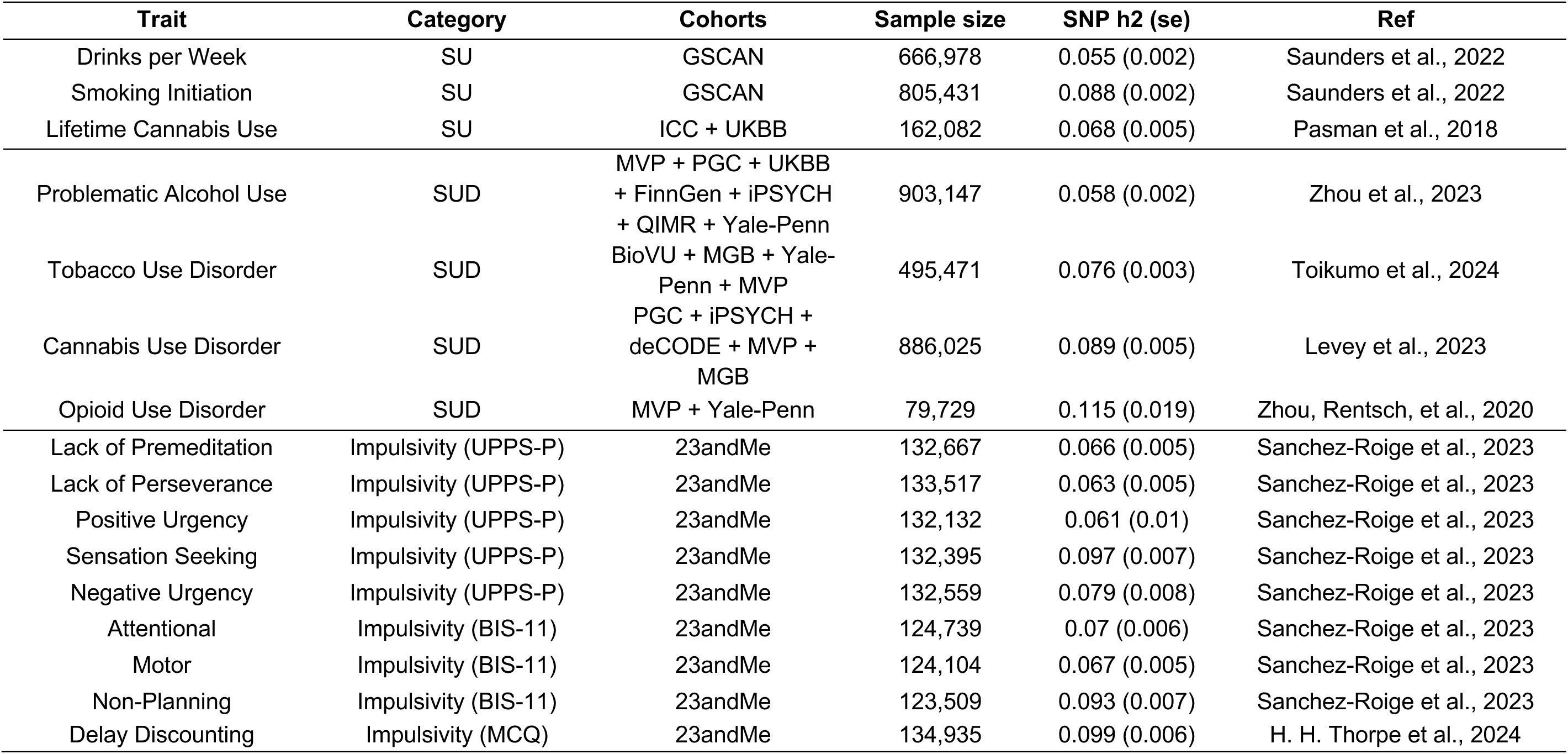
Summary of GWAS datasets for impulsivity, substance use and SUDs.

| Trait | Category | Cohorts | Sample size | SNP h2 (se) | Ref |
| --- | --- | --- | --- | --- | --- |
| Drinks per Week | SU | GSCAN | 666,978 | 0.055 (0.002) | Saunders et al., 2022 |
| Smoking Initiation | SU | GSCAN | 805,431 | 0.088 (0.002) | Saunders et al., 2022 |
| Lifetime Cannabis Use | SU | ICC + UKBB | 162,082 | 0.068 (0.005) | Pasman et al., 2018 |
| Problematic Alcohol Use | SUD | MVP + PGC + UKBB<br>+ FinnGen + iPSYCH<br>+ QIMR + Yale-Penn | 903,147 | 0.058 (0.002) | Zhou et al., 2023 |
| Tobacco Use Disorder | SUD | BioVU + MGB + Yale-Penn + MVP | 495,471 | 0.076 (0.003) | Toikumo et al., 2024 |
| Cannabis Use Disorder | SUD | PGC + iPSYCH +<br>deCODE + MVP +<br>MGB | 886,025 | 0.089 (0.005) | Levey et al., 2023 |
| Opioid Use Disorder | SUD | MVP + Yale-Penn | 79,729 | 0.115 (0.019) | Zhou, Rentsch, et al., 2020 |
| Lack of Premeditation | Impulsivity (UPPS-P) | 23andMe | 132,667 | 0.066 (0.005) | Sanchez-Roige et al., 2023 |
| Lack of Perseverance | Impulsivity (UPPS-P) | 23andMe | 133,517 | 0.063 (0.005) | Sanchez-Roige et al., 2023 |
| Positive Urgency | Impulsivity (UPPS-P) | 23andMe | 132,132 | 0.061 (0.01) | Sanchez-Roige et al., 2023 |
| Sensation Seeking | Impulsivity (UPPS-P) | 23andMe | 132,395 | 0.097 (0.007) | Sanchez-Roige et al., 2023 |
| Negative Urgency | Impulsivity (UPPS-P) | 23andMe | 132,559 | 0.079 (0.008) | Sanchez-Roige et al., 2023 |
| Attentional | Impulsivity (BIS-11) | 23andMe | 124,739 | 0.07 (0.006) | Sanchez-Roige et al., 2023 |
| Motor | Impulsivity (BIS-11) | 23andMe | 124,104 | 0.067 (0.005) | Sanchez-Roige et al., 2023 |
| Non-Planning | Impulsivity (BIS-11) | 23andMe | 123,509 | 0.093 (0.007) | Sanchez-Roige et al., 2023 |
| Delay Discounting | Impulsivity (MCQ) | 23andMe | 134,935 | 0.099 (0.006) | H. H. Thorpe et al., 2024 |

#### Impulsivity

We used summary statistics from our latest impulsivity GWAS (Sanchez-Roige et al., 2023). These included five measures from the UPPS-P Impulsive Behavior Scale (Cyders et al., 2014; Whiteside et al., 2005) and three from the BIS scales (Barratt, 1994) in a cohort comprising up to 133,517 consented research participants from 23andMe, Inc. These data sets have been extensively described elsewhere (Sanchez-Roige et al., 2023).

#### Delay discounting

We used summary statistics from our latest GWAS of delay discounting (H. H. A. Thorpe et al., 2024), which included 134,935 consented research participants from 23andMe, Inc. Participants completed the 27-item MCQ (Kirby et al., 1999). More information about this dataset can be found in H. H. Thorpe et al., (2024).

#### Substance use

We used summary statistics from three normative substance use GWAS: drinks per week (N = 666,978; Saunders et al., 2022), smoking initiation (N = 805,431; Saunders et al., 2022) and lifetime cannabis use (N = 184,765; Pasman et al., 2018).

#### Substance use disorders

We used summary statistics from four SUDs GWAS: problematic alcohol use (PAU) (N = 903,147; Zhou et al., 2023)), tobacco use disorder (TUD) (N = 495,471; Toikumo et al., 2024), cannabis use disorder (CUD) (N = 886,025; Levey et al., 2023) and opioid use disorder (OUD) (N = 79,729; Zhou, Rentsch, et al., 2020).

### Data analysis

All analyses were conducted in R (Version 4.1.1; R CoreTeam, 2022). We used the GenomicSEM package (Version 0.0.4; Grotzinger et al., 2019), which applies SEM methods to GWAS summary statistics. Genomic SEM leverages Linkage Disequilibrium Score Regression (LDSC) (Bulik-Sullivan et al., 2015) to generate a genetic correlation matrix between traits from GWAS summary statistics. Genomic SEM adjusts for potential sample overlap by estimating a sampling covariance matrix that indexes the precision of the estimates as well as the extent to which the sampling dependencies of the estimates are associated (Grotzinger et al., 2019).

Structural equation models are fit to the data using Genomic SEM, which draws on functionality from the *lavaan* R package (Rosseel, 2012). We used standard European reference panels and parameter settings, and the diagonally weighted least squares (DWLS) estimation method. Model fit was determined based on the comparative fit index (CFI) and the standardized root mean square residual (SRMR). Good-fitting models are expected to have CFI higher than .95 (.90 for acceptable fit), SRMR smaller than .08, and smaller AIC values than competing nested models (Hu & Bentler, 1998). Significance of individual parameter estimates was established with 95% confidence intervals (CIs) and FDR corrected *p* value.

### Model-fitting approach

First, we separately fit confirmatory factor models of impulsivity and substance use-related measures. For impulsivity, we fit a six-factor model, using GWAS data from the UPPS-P (five traits) and the BIS (three traits) subscales and delay discounting (**Table 1**), based on our prior work (Gustavson et al., 2020, 2024). This model included the three BIS subscales and the UPPS-P lack of premeditation subscale as indices of a lack of premeditation factor (**Figure 2**). Next, we fit a two-factor model comprising the SU and SUD factors, using GWAS data from seven substance use-related measures, based on prior work by Hatoum et al., (2022) and Karlsson Linnér et al., (2021). Notably, our SU factor is composed of indices of lifetime use as well as quantity/frequency of use. Despite the lack of GWAS for lifetime alcohol use, the high genetic correlation between alcohol quantity/frequency and smoking/cannabis lifetime use measures (**Figure 1**) suggests that these phenotypes could potentially be indicative of a single latent factor. Our SUD factor replicated the addiction-risk-factor model by Hatoum et al., (2022, 2023), replacing problematic tobacco use for TUD (Toikumo et al., 2024), which was not available at the time of their analyses. Finally, we constructed a common factor model in which all factors (six impulsivity factors, SU factor, SUD factor) were fitted simultaneously. We fit two versions of this model: (a) a correlated factors model where we estimated the genetic correlations between all impulsivity, SU, and SUD factors, and (b) a multiple regression mode, where impulsivity was regressed on SU and SUD factors, with the latent factors treated as correlated outcomes. Within the correlated factors model, to test whether impulsivity facets were differentially correlated with the SU and SUD factors, we included a computed variable (i.e., a defined parameter) that captured the difference between the SU and SUD estimates. We used the associated *p* value to test this difference. The multiple regression model was used to quantify whether genetic associations between impulsivity and SU were independent of genetic influences on SUD (and vice-versa). To account for multiple testing, we applied an FDR correction to all *p* values.

**Figure 1.**
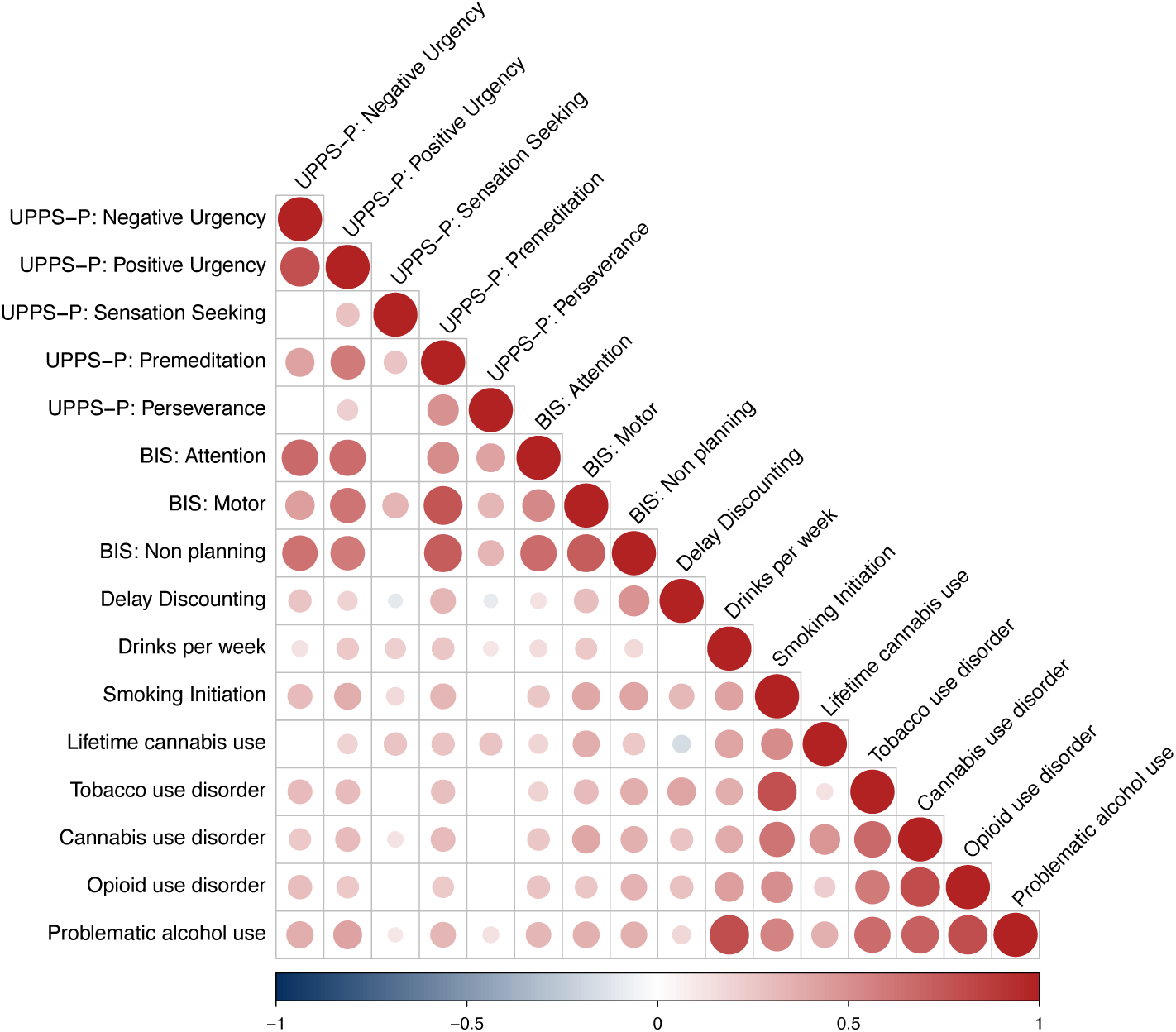
Genetic correlation (*r_g_*) matrix between all study variables. The size and color of the circle indicates the strength of the correlation. Non-significant genetic correlations are left blank. *r_g_* and 95% confidence intervals can be found in **Supplementary Table 1**. UPPS-S = Impulsive Behavior Scale; BIS = Barratt Impulsiveness Scale.

**Figure 2.**
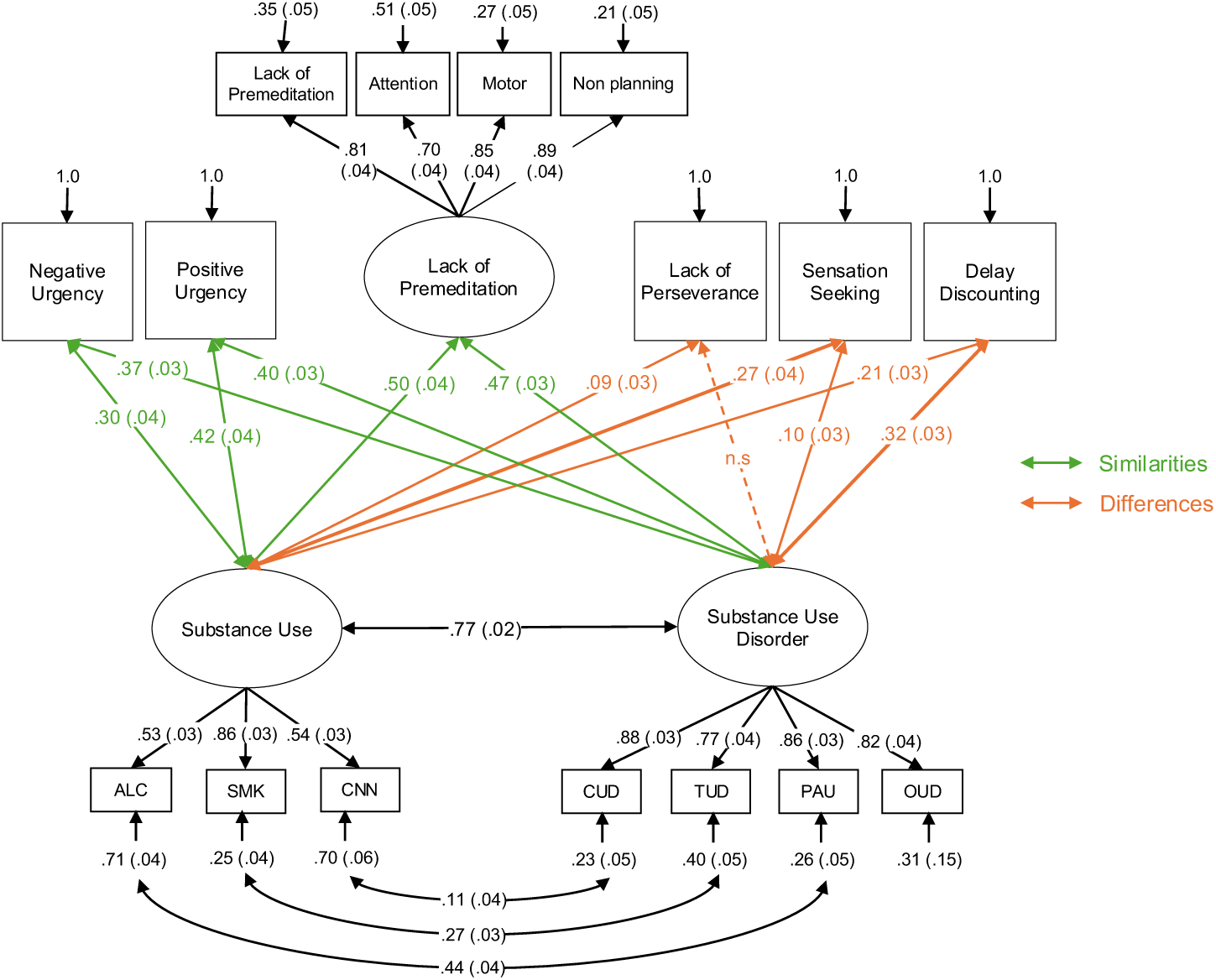
Path diagram for the final model of impulsivity, substance use (SU) and substance use disorder (SUD) traits. Observed indicators are represented by squares and latent factors are represented by circles. Single headed arrows indicate factor loading, and double headed arrows indicate correlations. Orange arrows represent genetic correlations significantly different between impulsivity and SU versus SUD, and green arrows represent genetic correlations of similar magnitude between impulsivity and SU or SUD. All values indicate standardized parameters estimates. Confidence intervals and *p* values can be found in **Supplementary Table 2**.

## Data availability

Summary statistics for the SU and SUD GWAS were publicly available. Data from 23andMe are available upon request (see https://research.23andme.com/dataset-access/). The R data files containing the Genomic SEM matrices and an R markdown with the analysis scripts and corresponding outputs are available at the following link https://osf.io/ba8dy/?view_only=acacd8c2eae844e88b8b4510a7d59973, which allows for replication and analyses of competing models without obtaining the source data.

## Results

Figure 1 shows the genetic correlation matrix (*r_g_*) among all study variables. Genetic correlations between impulsivity facets and substance use involvement traits were broadly significant and positive, ranging from *r_g_ =* 0.11 to *r_g_ =* 0.80, with a few exceptions (**Supplementary Table 1**). Delay discounting was negatively genetically correlated with sensation seeking, lack of perseverance and lifetime cannabis use (*r_g_ range* = −0.1 to *-*0.16).

The six-factor impulsivity model, which was based on prior work from (Gustavson et al., 2020), showed acceptable fit (χ^2^(17) = 441.49, CFI = .938, SRMR = .074) (Figure 2). This model included five single-item factors (i.e., negative urgency, positive urgency, lack of perseverance, sensation seeking and delay discounting), and one common factor comprising the three BIS subscales and lack of premeditation. The two-factor substance use model, which captured the shared genetic variation of drinks per week, smoking initiation and lifetime cannabis use into a SU factor, and the shared genetic variation of PAU, TUD, CUD and OUD into a SUD factor, also showed an acceptable fit (χ^2^(10) = 98.57, CFI = .985, SRMR = .056). As expected, the SU and SUD factors exhibited a strong positive genetic correlation, indicating significant shared genetic influences (*r_G_* = .77, 95% CI = [0.73, 0.81]). Because we expected that measures related to the same substance (e.g., drinks per week and PAU) would be especially correlated with one another, we also included residual correlations between these measures in our model. The residual correlation between drinks per week and PAU was moderate (*r_G_* = .44, 95% CI = [0.36, 0.52]), while the residual correlations between smoking initiation and TUD, and lifetime cannabis use and CUD, were relatively low (*r_G_* = .27, 95% CI = [0.21, 0.34] and .11, 95% CI = [0.04, 0.19], respectively).

Our final common factor model, displayed in Figure 2, combined the six impulsivity, SU and SUD factors (χ^2^(78) = 2392.55, CFI = .939, SRMR = .072). The SU and SUD factors were most strongly genetically correlated with lack of premeditation (SU *r_G_* = .50, 95% CI = [0.42, 0.58]; SUD *r_G_* = .47, 95% CI = [0.4, 0.53]), and least strongly genetically correlated with lack of perseverance (SU *r_G_* = .09, 95% CI = [0.03, 0.16]; SUD *r_G_* = .04, 95% CI = [-0.02, 0.1], *p* = n.s.). Sensation seeking was more strongly genetically correlated with the SU factor than with the SUD factor (*r_G_* = .27, 95% CI = [0.19, 0.34] vs. *r_G_* = .10, 95% CI = [0.04, 0.17], respectively, *p* = 4.17e-05). On the contrary, delay discounting was more strongly genetically correlated with the SUD factor than with the SU factor (*r_G_* = .32, 95% CI = [0.27, 0.38] vs. *r_G_* = .21, 95% CI = [0.15, 0.28], respectively, *p* = 2.41e-03). For all other impulsivity facets, genetic correlations with the SU and SUD factors were not significantly different from one another.

Given the strong genetic correlation between the SU and SUD factors, we performed a specificity analysis via a multiple regression model to test whether these factors were independently associated with impulsivity facets. This final model had the same fit as the correlated factor model (from Figure 2), and the results are described in Figure 3 and **Supplementary Table 3**. We observed distinct patterns of associations for most of the impulsivity facets with the SU and SUD factors. Controlling for SUD, the SU factor was positively genetically associated with positive urgency (beta (β) = 0.28, 95% CI = [0.1, 0.45], *p* = 2.94e-03), lack of premeditation (β = 0.39, 95% CI = [0.18, 0.61], *p* = 4.80e-04), lack of perseverance (β = 0.16, 95% CI = [0.01, 0.31], *p* = 4.88e-02), and sensation seeking (β = 0.46, 95% CI = [0.3, 0.62], *p* = 8.65e-08). Controlling for SU, the SUD factor was positively genetically associated with negative urgency (β = 0.34, 95% CI = [0.2, 0.48], *p* = 2.53e-06), positive urgency (β = 0.19, 95% CI = [0.04, 0.34], *p* = 1.64e-02), lack of premeditation (β = 0.24, 95% CI = [0.06, 0.42], *p* = 1.15e-02) and delay discounting (β = 0.39, 95% CI = [0.27, 0.51], *p* = 5.68e-10), and negatively genetically associated with sensation seeking (β = −0.25, 95% CI = [-0.4, −0.1], *p* = 1.98e-03).

**Figure 3.**
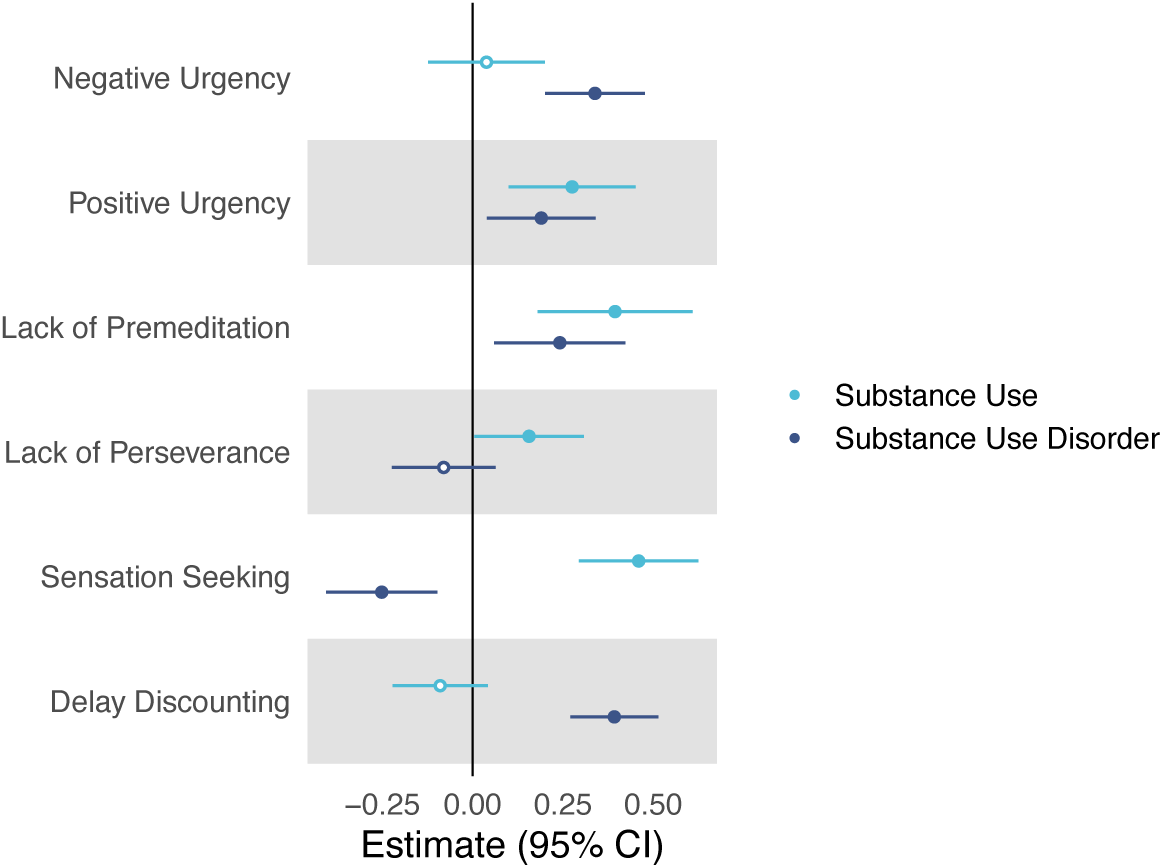
Standardized genetic associations between SU and SUD (controlling for one another), and the six impulsivity factors. Estimates, 95% confidence intervals and *p* values for these regressions can be found in **Supplementary Table 3**. Significant associations are indicated with a filled-in circle.

## Discussion

A deeper understanding of the relationship between impulsivity facets and stages of substance use vulnerability can facilitate the development of better prevention, diagnosis and treatment strategies for SUDs. In this study, we used GWAS data and Genomic SEM techniques to model the genetic architecture of nine impulsivity facets and seven measures related to substance use and SUDs. Virtually all facets of impulsivity were genetically correlated with both the SU and SUD factors, but the magnitude of these correlations varied across different impulsivity facets. Our findings provide evidence that specific impulsivity facets confer risk for distinct stages of substance use involvement and emphasize the need to consider the multi-dimensional nature of impulsivity in SUD-related research.

As expected, the SU and SUD factors were strongly genetically correlated but not at unity, consistent with our prior work (Hatoum et al., 2022, 2023). This supports the idea that while normative use and SUDs are related, they have different genetic architectures, which may also contribute to differences in their relationships with other traits or behaviors (Dick et al., 2011; Kranzler et al., 2019; Levey et al., 2023; Mallard et al., 2022; Mallard & Sanchez-Roige, 2021; Polimanti et al., 2020; Sanchez-Roige et al., 2022; Sanchez-Roige, Palmer, et al., 2019). For example, some impulsivity facets were most strongly linked to aspects of normative use. In particular, sensation seeking showed a stronger genetic correlation with the SU than the SUD factor. Delay discounting, however, showed a stronger genetic correlation with the SUD than the SU factor. Meanwhile, negative and positive urgency, lack of premeditation and lack of perseverance showed similar genetic correlations with both the SU and SUD factors. Most of these results held in our regression model, which accounted for the genetic overlap between the SU and SUD. Our findings further highlight impulsivity facets as independent phenotypic and genetic constructs, in some cases only moderately correlated with each other (Gustavson et al., 2019, 2020; Sanchez-Roige et al., 2023; H. H. Thorpe et al., 2024), that capture specific biological processes relevant to normative and disordered substance use (Miller & Gizer, 2024; Mitchell, 2019; Poulton & Hester, 2020).

Sensation seeking, defined as the tendency to engage in highly stimulating behaviors, was more strongly genetically correlated with the SU factor compared to the SUD factor, which was further supported by our regression model. Indeed, among all impulsivity facets, sensation seeking exhibited the strongest genetic association with the SU factor. Our results contribute to a substantial body of evidence demonstrating that sensation seeking is primarily associated with aspects of initiation and experimental substance use (Birkley & Smith, 2011; Fischer et al., 2008; Griffin et al., 2018; Hildebrandt et al., 2021; Stautz & Cooper, 2013). After controlling for SU, we also observed a significant negative genetic association between sensation seeking and the SUD factor, which aligns with previous phenotypic (Courtney et al., 2012) and genetic (Miller & Gizer, 2024) studies and suggests that the influence of sensation seeking on SUDs may primarily manifest through increased normative substance use.

Lack of perseverance, which reflects the cognitive difficulties of maintaining effort over an extended period of time, showed a unique, but weak, genetic correlation with the SU factor, but not with the SUD factor. This finding remained consistent in our regression model, and aligns with prior phenotypic studies (Fischer & Smith, 2008; Hildebrandt et al., 2021; Kearns et al., 2022; Magid & Colder, 2007; Stamates & Lau-Barraco, 2017) and our prior GWAS (Sanchez-Roige et al., 2023; Sanchez-Roige, Fontanillas, et al., 2019), suggesting that lack of perseverance may have a distinct genetic architecture and weaker genetic associations with substance use-related outcomes compared to other impulsivity facets (Gustavson et al., 2020, 2024).

Conversely, delay discounting showed a significantly stronger genetic correlation with the SUD factor compared to the SU factor, and as observed in the regression model, the strongest genetic association with the SUD factor among all impulsivity facets. This finding aligns with clinical- and population-based phenotypic studies, where devaluation of delayed rewards appears most relevant to problematic substance use over normative use (Courtney et al., 2012; Fröhner et al., 2022; Kräplin et al., 2020; P. Murphy & Garavan, 2011; Stamates & Lau-Barraco, 2017).

Positive and negative urgency showed equal genetic correlations with the SU and SUD factors, however, this was not consistent in our regression model. While positive urgency was independently associated with both the SU and SUD factors, negative urgency was uniquely associated with the SUD factor. This specificity aligns with theoretical models proposing that positive urgency, driven by enhanced sensitivity to rewards, may promote both initial experimentation and escalation to substance misuse, whereas negative urgency, reflecting difficulties regulating responses to negative emotions, may specifically contribute to compulsive use via a process of negative reinforcement during withdrawal (Cyders & Smith, 2008; Koob & Le Moal, 2001; Koob & Volkow, 2010; Smith & Cyders, 2016). Prior epidemiological evidence aligns with this specificity. Positive urgency has been previously associated with both normative (Kaiser et al., 2016; Tomko et al., 2016; Zapolski et al., 2009), and problematic substance use (Coskunpinar et al., 2013; Stautz & Cooper, 2014; Tran et al., 2018), whereas negative urgency has been primarily implicated in SUDs severity and related problems (Adams et al., 2012; Coskunpinar et al., 2013; Fischer & Smith, 2008; Hildebrandt et al., 2021; Kearns et al., 2022; Magid & Colder, 2007; C. Murphy & Mackillop, 2012; Stamates & Lau-Barraco, 2017; Stautz & Cooper, 2013; Tomko et al., 2016; Verdejo-Garcia & Albein-Urios, 2021), supporting our results. Despite the high genetic correlation between positive and negative urgency, our results suggest some distinct genetic relationships with stages of substance use vulnerability. This finding reinforces the notion that these impulsivity facets should be considered separate constructs (Billieux et al., 2021; Carver & Johnson, 2018).

Lastly, lack of premeditation, a facet characterized by low executive control and difficulties with prospective thinking and planning, showed the strongest genetic correlation with both the SU and SUD factors. Our regression model suggests that lack of premeditation has independent genetic contributions to both the SU, which aligns with prior phenotypic studies (Fischer & Smith, 2008; Griffin & Trull, 2021; McCabe et al., 2015; Shin et al., 2012, 2013), as well as the SUD factor, where prior findings have been mixed (Fischer & Smith, 2008; Hershberger et al., 2017; Hildebrandt et al., 2021; Magid & Colder, 2007; McCabe et al., 2015; Shin et al., 2012).

Our findings should be interpreted in the context of the following limitations. First, GWAS were only conducted in individuals with European genetic similarity; while we have no specific reason to believe these findings are specific to Europeans, future analyses should diversify genetic analyses as larger non-European samples become available. Additionally, all impulsivity GWAS were conducted in a cohort of 23andMe participants that were generally older and had higher socioeconomic status than the general population (Sanchez-Roige et al., 2023). These factors may introduce biases due to gene-environment correlations (Abdellaoui et al., 2022; H. H. A. Thorpe et al., 2024) and reduce the generalizability of the results. Furthermore, Genomic SEM allows modeling the genetic architecture between the traits studied, but it does not infer causality or directionality. Impulsivity facets can represent both risk factors as well as consequences of substance use (Kaiser et al., 2016; Verdejo-García et al., 2008), as seen in longitudinal, family-based and clinical studies where individuals with increased impulsive behaviors have higher susceptibility to substance use and SUDs (Verdejo-Garcia & Albein-Urios, 2021). In addition, chronic substance use can lead to brain alterations that can further exacerbate impulsivity (de Wit, 2009). However, the 23andMe cohort used in the impulsivity GWAS data reported relatively low levels of drug use, suggesting that impulsivity due to recent drug use is unlikely to be a major confounder in our study (Sanchez-Roige et al., 2023). Furthermore, the SU and SUD factors exhibited a strong genetic correlation, complicating the interpretation of the regression model due to collinearity issues. Despite this challenge, it remains crucial to distinguish between normative use and SUDs, even if the unique variance is small. This differentiation will inform more effective prevention and intervention strategies aimed at preventing normative SU from progressing to SUDs. Finally, it is noteworthy that some of our results differ from those reported in previous phenotypic studies. However, genetic studies and phenotypic observations can sometimes diverge due to factors like gene-environment interplay, sampling error and confounding factors not accounted for in the analysis (e.g. socioeconomic status, peer influences, or availability of substances) (Sodini et al., 2018).

In conclusion, our study provides novel insights into the complex genetic relationships between distinct facets of impulsivity and different stages of substance use involvement. Our findings highlight both similarities and differences on the genetic contributions of various impulsivity domains to aspects of normative substance use and SUDs. These results reinforce the importance of studying impulsivity as multidimensional constructs when trying to determine their role in SUDs vulnerability. Future work should further explore the underlying biological mechanisms of individual facets of impulsivity, many of which are amenable to studies in non-human animals. The insights from this study can also be leveraged to inform targeted prevention and intervention efforts tailored to individual impulsivity profiles and substance use risk.

## Acknowledgements

We would like to thank the research participants and employees of 23andMe for making this work possible.

The following members of the 23andMe Research Team contributed to this study:

Stella Aslibekyan, Adam Auton, Elizabeth Babalola, Robert K. Bell, Jessica Bielenberg, Ninad S. Chaudhary, Zayn Cochinwala, Sayantan Das, Emily DelloRusso, Payam Dibaeinia, Sarah L. Elson, Nicholas Eriksson, Chris Eijsbouts, Teresa Filshtein, Pierre Fontanillas, Davide Foletti, Will Freyman, Zach Fuller, Julie M. Granka, Chris German, Éadaoin Harney, Alejandro Hernandez, Barry Hicks, David A. Hinds, M. Reza Jabalameli, Ethan M. Jewett, Yunxuan Jiang, Sotiris Karagounis, Lucy Kaufmann, Matt Kmiecik, Katelyn Kukar, Alan Kwong, Keng-Han Lin, Yanyu Liang, Bianca A. Llamas, Aly Khan, Steven J. Micheletti, Matthew H. McIntyre, Meghan E. Moreno, Priyanka Nandakumar, Dominique T. Nguyen, Jared O’Connell, Steve Pitts, G. David Poznik, Alexandra Reynoso, Shubham Saini, Morgan Schumacher, Leah Selcer, Anjali J. Shastri, Jingchunzi Shi, Suyash Shringarpure, Keaton Stagaman, Teague Sterling, Qiaojuan Jane Su, Joyce Y. Tung, Susana A. Tat, Vinh Tran, Xin Wang, Wei Wang, Catherine H. Weldon, Amy L. Williams, Peter Wilton.

